# Elevation of Neurodegenerative Serum Biomarkers among Hospitalized COVID-19 Patients

**DOI:** 10.1101/2021.09.01.21262985

**Authors:** Jennifer A. Frontera, Allal Boutajangout, Arjun V. Masurkar, Rebecca A. Betensky, Yulin Ge, Alok Vedvyas, Ludovic Debure, Andre Moreira, Ariane Lewis, Joshua Huang, Sujata Thawani, Laura Balcer, Steven Galetta, Thomas Wisniewski

## Abstract

**INTRODUCTION:** Older adults hospitalized with COVID-19 are susceptible to neurological complications, particularly encephalopathy, which may reflect age-related neurodegenerative processes.

**METHODS:** Serum total tau, ptau-181, GFAP, NFL, UCHL1, and amyloid-beta(Aβ-40,42) were measured in hospitalized COVID-19 patients without a history of dementia, and compared among patients with or without encephalopathy, in-hospital death versus survival, and discharge home versus other dispositions using multivariable Cox proportional hazards regression analyses.

**RESULTS:** Among 251 patients, admission serum ptau-181 and UCHL1 were significantly elevated in patients with encephalopathy (both P<0.05) and total tau, GFAP, and NFL were significantly lower in those discharged home(all P<0.05). These markers correlated significantly with severity of COVID illness. NFL, GFAP and UCH-L1 were significantly higher in hospitalized COVID patients than in non-COVID controls with mild cognitive impairment or Alzheimer’s disease(AD).

**DISCUSSION:** Age-related neurodegenerative biomarkers were elevated to levels observed in AD and associated with encephalopathy and worse outcomes among hospitalized COVID-19 patients.

## INTRODUCTION

Neurological complications, particularly encephalopathy, are common among hospitalized COVID-19 patients (1, 2), and long-term cognitive abnormalities persist in nearly 50% of hospital survivors (3). However, the mechanisms underpinning cognitive dysfunction in acute and post-acute COVID-19 patients are not well understood. One possibility is that protracted hypoxia and the hyperinflammatory state encountered in acute, severe COVID-19 may lead to neuronal and glial cell injury, which could be measured in the blood using sensitive digital ELISA or single molecule array technology (SIMOA).

In this study, we hypothesized that blood biomarkers of neuronal and glial injury would be elevated in hospitalized COVID-19 patients with clinical evidence of new onset of cognitive dysfunction (specifically toxic-metabolic encephalopathy [TME]), and that elevated neurodegenerative biomarkers would be associated with a higher risk of in-hospital death and reduced rates of discharge home. We further aimed to compare neurodegenerative biomarker levels in hospitalized COVID patients to non-COVID controls with varying degrees of cognitive impairment (normal, mild cognitive impairment [MCI] or Alzheimer’s Disease [AD]) to gauge their degree of brain injury.

We chose to assess neurofilament light chain (NFL), a cytoskeletal intermediate filament protein integral to axons in the central and peripheral nervous system, which has been noted to be elevated in patients with COVID-19(4-9), as well as glial fibrillary acidic protein (GFAP), which is specific indicator of glial/astrocyte injury, and ubiquitin carboxy-terminal hydrolase L1 (UCHL1) and total tau, which are a neuron-specific proteins. Plasma total tau, NFL and GFAP have also been reported to be elevated in AD (10-12). Additionally, we evaluated phosphorylated tau-181 (ptau181), and amyloid beta (Aβ)-40 and 42, which are more specific biomarkers for Alzheimer’s type pathology (10, 13-15). Notably, UCH-L1(16, 17), GFAP(18), tau(18), and NFL(19, 20) are also elevated following blood brain barrier disruption, which has been documented in neuropathological studies of COVID-19 decedents(21) as well as in AD.

## METHODS

### Study Design and Patient Cohort

We conducted a retrospective analysis of COVID-19 patients who were prospectively enrolled in the Study of Neurologic and Psychiatric Events in Acute COVID-19 (SNaP Acute COVID) study (22), and had serum biospecimens collected and banked during their index hospitalization for COVID-19. Briefly, SNaP Acute COVID is a prospective study of consecutive COVID-19 patients hospitalized at four New York City area hospitals within the same hospital system between March 10, 2020 and May 20, 2020. Patients were prospectively evaluated by a team of neurologists for development of new neurological disorders during acute COVID-19 hospitalization. For our current study, inclusion criteria were: hospital admission, reverse-transcriptase-polymerase-chain-reaction (RT-PCR) positive SARS-CoV-2 infection from nasopharyngeal sampling, and consent to store blood biospecimens in the NYU Center for Biospecimen Research and Development biorepository for use in experimental analyses. All COVID-19 patients capable of consent were approached for blood banking at admission. Exclusion criteria were: negative or missing SARS-CoV-2 RT-PCR test, evaluation in an outpatient or emergency department setting only, history of dementia or cognitive impairment (including but not limited to: pre-existing diagnoses of MCI, AD, vascular dementia, Lewy body/Parkinson’s related dementia, progressive supranuclear palsy, multiple system atrophy, corticobasal degeneration, frontal-temporal dementia, normal pressure hydrocephalus or Creutzfeld-Jakob disease), and inadequate biospecimens available for analyses.

Control populations of non-COVID-19 subjects with blood samples banked prior to January 1, 2020 (prior to the first reported cases of SARS-CoV-2 infection in NYC) were selected from the NYU Alzheimer’s Disease Research Center (ADRC) Clinical Core cohort. Three control populations were included: cognitively normal (defined by normal Uniform Data Set version 3 [UDS-3] psychometric testing and Clinical Dementia Rating [CDR] score of 0), MCI (defined by abnormal UDS-3 psychometric testing and CDR = 0.5) and dementia due to Alzheimer’s disease (AD) (defined as abnormal UDS-3 psychometric testing, CDR ≥ 1, and clinical phenotype/biomarker suggestive of primary AD and not other dementia subtypes(23, 24)).

### Blood banking process

After obtaining consent, leftover blood samples drawn on hospital day 0 during index COVID-19 hospitalization were banked for future research. Serum samples were collected from COVID-19 patients in gold or red top tubes and kept at room temperature for 30-45 minutes after blood draw to assess for clot formation prior to centrifugation at 4°C 2000G for 10 minutes.

Non-COVID control plasma biospecimens were collected at the NYU ADRC in EDTA tubes and kept on wet ice until centrifugation at 4°C 2000G for 10 minutes. Both serum and plasma samples were aliquoted 0.200 mL into 1mL polypropylene tubes and stored in -80°C freezers equipped with 24h alarm systems for detecting temperature excursions. Because only plasma control specimens were available, we evaluated only NLF, GFAP and UCHL-1 levels for comparison with the COVID-19 samples. These three biomarkers been shown to have equivalent levels in serum and plasma samples, whereas significant differences have been identified in total tau and Aβ levels between serum and plasma(25, 26).

### Neurodegenerative biomarker analyses

Serum and plasma biomarker assays were conducted by the Biomarker Core of the NYU Alzheimer’s Disease Research Center using the Single Molecule Array Technology SR-X (SIMOA SR-X™ Analyzer, Quanterix Corporation, MA). Total tau (Tau), Neuro-Filament light (NFL), Glial fibrillary acidic protein (GFAP), and Ubiquitin Carboxy-Terminal Hydrolase L1 (UCH-L1) were measured using the Simoa® Neurology 4-plex A kit. The Simoa® pTau-181 Advantage Kit was used to measure ptau-181. Aβ-40 and 42 were measured using the Simoa® Neurology 3-plex A kit. Following the manufacturer recommendation for handling and analyzing serum samples, each Simoa kits run included: an 8-point calibration curve for each marker and 2 internal controls.

To avoid batching effects, experiments were predesigned including a similar number of individuals from all study groups once sufficient samples were collected. Investigators running the experiments were blinded to study group assignments. The samples were thawed once, in ice, before each run, and centrifuged 5 minutes at 10,000g before being manually diluted 1:4 in the 96 wells plate with the appropriate buffer included in Simoa® kits. Each sample was run in duplicate and the average value of both runs was used for analyses.

### Other laboratory data

Blood inflammatory markers collected as part of clinical care during the hospital encounter were abstracted from the medical record including interleukin-6 (IL-6), C-reactive peptide (CRP), D-Dimer and ferritin levels. The values obtained at hospital day 0 were used for analyses.

### Neurological Diagnoses and Severity of Illness Scales

Neurological diagnoses (including TME, hypoxic-ischemic encephalopathy, stroke [ischemic or hemorrhagic], seizure, neuropathy, myopathy, movement disorder, encephalitis/meningitis, myelopathy, myelitis) followed established criteria and were coded for COVID-19 patients found to have a *new* neurological complication (excluding recrudescence or worsening of old neurological deficits) as diagnosed by in-hospital neurology teams. TME was coded for patients with new changes in mental status in the absence of focal neurological deficits (except in cases of hypo/hyperglycemia), clinical or electrographic seizures or primary structural brain disease. For patients who had received sedating medications, an adequate washout (4-5 half-lives) was required for mental status assessment prior to diagnosis with TME. Patients with altered mental status due to another acute neurological diagnosis that could account for the observed exam findings (e.g. stroke, seizure, traumatic brain injury)(27) or abnormal mental status due to sedative medications were excluded from the TME diagnostic category. Subsidiary review of all neurological diagnoses was performed by relevant neurological subspecialists on the study team (e.g. stroke, neurocritical care, epilepsy sub-specialists). Patients could be coded for more than one neurological complication.

Demographic data, past medical history, clinical course and hospital outcomes (mortality rates discharge disposition, ventilator days and hospital length of stay) were collected. Severity of illness during hospitalization was assessed using the worst recorded Sequential Organ Failure Assessment (SOFA) score. Past neurological history included: history of ischemic or hemorrhagic stroke, hydrocephalus, brain tumor, headache, seizure, traumatic brain injury, neuropathy, myasthenia gravis, multiple sclerosis, or movement disorder.

### Study Outcomes

The primary outcomes were serum levels of Tau, pTau181, NFL, GFAP, UCHL1, Aβ-40, Aβ-42, the ratio of Aβ-42/Aβ-40 and the ratio of pTau181/Aβ-42 compared between hospitalized COVID-19 patients who: 1) developed TME versus those who did not, 2) died in-hospital/discharged to hospice versus those who survived to discharge, and 3) were discharged home versus other discharge dispositions (in-hospital death/hospice, discharge to a nursing home, long-term acute care facility, acute or subacute rehabilitation facility). Secondary outcomes included the comparison of serum biomarker levels among COVID-19 patients to plasma biomarker levels of NFL, GFAP and UCHL1 in non-COVID controls with normal cognition, MCI or AD.

### Standard Protocol Approvals and Patient Consents

This study was approved by the NYU Grossman School of Medicine Institutional Review Board. All patients or their surrogates provided consent for participation in blood banking.

### Statistical Analyses

The sample size was calculated based on detection of a 25% difference in biomarker levels between patients with or without TME during hospitalization, which has a prevalence of 30% in our patient sample (e.g. 1 case to 3 controls). Assuming a power of 80% and an alpha=0.05, 42 TME cases and 126 non-TME patients (total N=168) would be needed to detect a 25% difference in NFL, 62 TME cases and 186 non-TME patients (total N=248) would be needed to detect a 25% difference in UCHL-1, 38 TME cases and 1114 non-TME patients (total N=152) would be needed to detect a 25% difference in GFAP, 104 TME cases and 312 non-TME patients (total N=416) would be needed to detect a 25% difference in total tau levels, 33 TME cases and 99 non-TME patients (total N=132) would be needed to detect a 25% difference in Aß-40 levels, and 40 TME cases and 120 non-TME patients (total N=160) would be needed to detect a 25% difference in Aβ-42 levels.

Neurodegenerative and inflammatory laboratory values were reported as median and interquartile ranges (IQR). Specialized nonparametric U-statistics were used to compare biomarker levels between patients with and without TME, death, or discharge home. This statistic calculates pairwise rankings for comparable pairs, and accounts for the time needed to have the opportunity to be diagnosed with a specific outcome prior to the occurrence of a competing event. This statistic reduces to the Mann Whitney U-statistic when there is no adjustment for relative timing of diagnosis and length of hospital stay. We used the simple bootstrap (500 repetitions) to calculate the variances of the U-statistics. Biomarker levels were compared between COVID-19 patients, and non-COVID cognitively normal, MCI and AD patients using Mann-Whitney U non-parametric tests. Correlations between biomarker levels and demographics, severity of illness measures and inflammatory laboratory values were assessed using 2-tailed Spearman’s rank correlation coefficients.

Using cause-specific multivariable Cox proportional hazard regression models, we fit the event times of: 1) diagnosis of TME; 2) in-hospital death/hospice discharge and; 3) discharge to home. For the outcome of TME, in-hospital death or hospital discharge (home, skilled nursing facility, acute or subacute rehab or long term acute care hospital [LTACH]) were treated as censoring events; for the outcome of in-hospital death, any discharge disposition other than death or discharge to hospice was treated as a censoring event; and for the outcome of discharge home, we treated death or discharge to hospice, a skilled nursing facility, acute or subacute rehab or long term acute care hospital [LTACH] as censoring events. No patients remained under observation in the hospital at the end of follow-up for this study. All models were adjusted for confounders including age, sex, race, history of neurological disease, admission sequential organ failure assessment (SOFA) score, and admission oxygen saturation. Covariates were selected based on known predictors of in-hospital death, poor discharge disposition, biological plausibility and bivariate associations within our own data(1, 3, 22). Analyses were conducted using IBM SPSS Statistics for Mac V25 (IBM Corp., Armonk, NY) and R studio V1.1.456.

## RESULTS

A total of 302 patients from the SNaP Acute COVID cohort had banked serum specimens available for analysis. After excluding 51 patients with a history of dementia or cognitive impairment, 251 COVID-19 patients were included in analysis (Figure 1). Due to limited sample availability,, the number of patients tested for each biomarker varied: NFL, GFAP, and UCHL1 were assayed in N=246, Tau in N=241, pTau181 in N=157, Aβ-40 in N=146 and Aβ-42 in N=120. A total of 161 controls underwent neurodegenerative biomarker testing (N=54 cognitively normal, N=54 MCI, N=53 AD).

**Figure 1.**
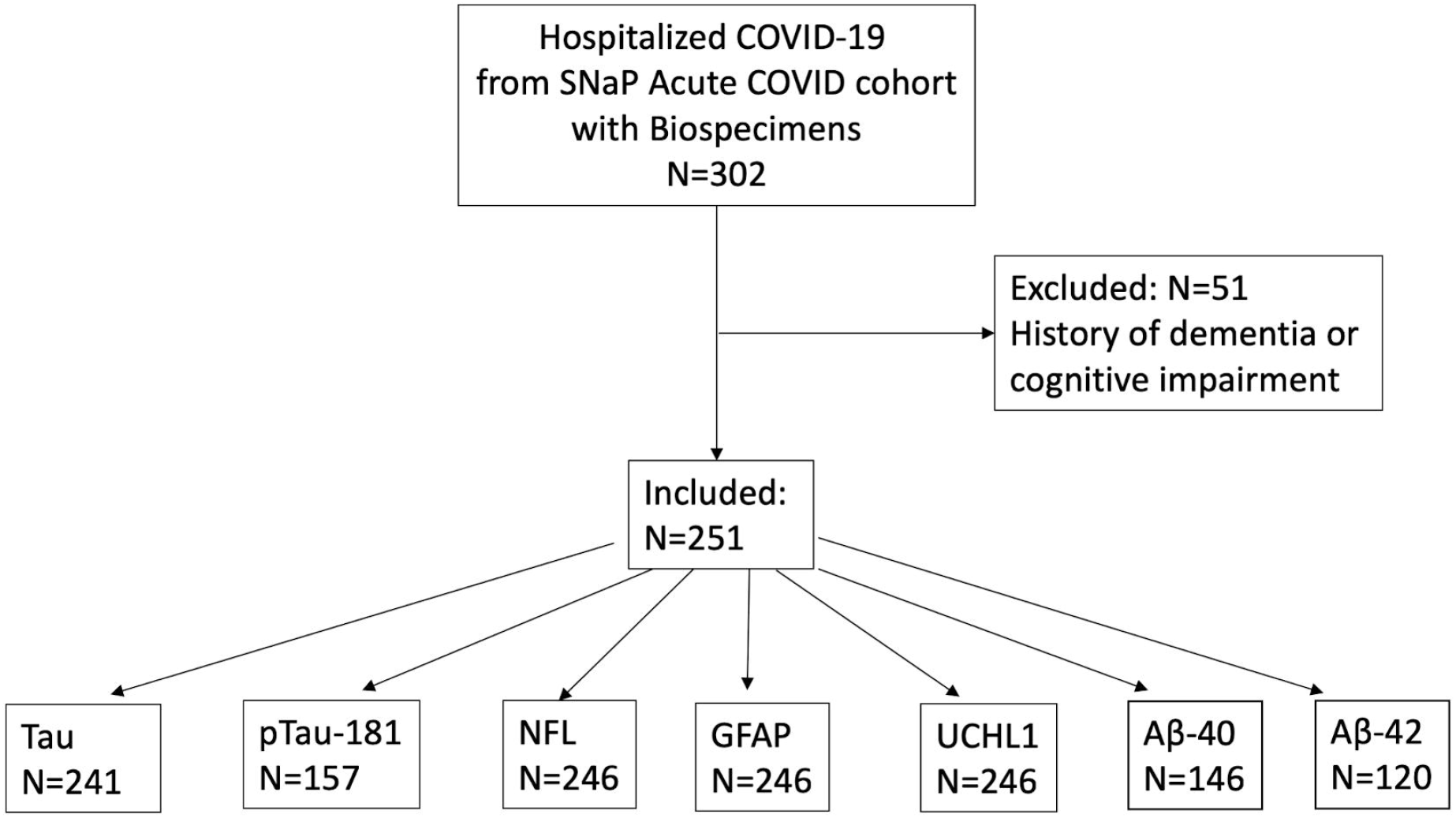
Flow chart of patient inclusion. SNaP Acute COVID= Study of Neurologic and Psychiatric Events in Acute COVID-19; pTau=phosphorylated Tau; NFL=neurofilament light chain; GFAP=glial fibrillary acidic protein; UCHL1=ubiquitin carboxyl-terminal hydrolase isozyme L1; Aß= amyloid beta.

The median age of COVID-19 patients was 71 years (IQR 60-83) and 63% were male, compared to 70 years (IQR 65-76) and 35% male among non-COVID cognitively normal controls (P=0.997 for age; P=0.001 for sex), 77 years (range 70-86) and 20% male among non-COVID MCI controls (P=0.002 for age; P<0.001 for sex), and 82 years (range 73-89) and 40% male among non-COVID AD patients (P<0.001 for age; P=0.002 for sex).

Among COVID-19 patients, 31% required mechanical ventilation, 25% died in-hospital and 53% were discharged home (Table 1). New neurological events during hospitalization occurred in 120/251 (48%) of patients with the most common diagnoses being TME in 75/120 (63%) and hypoxic/ischemic brain injury in 55/120 (46%). Among patients diagnosed with TME during their hospital stay, the median time from admission to diagnosis of TME was 0 days (IQR 0-3 days), the median time to death among those who died in-hospital was 11 days (IQR 6-22 days), and the median hospital length of stay was 10 days (IQR 6-20 days).

**Table1.**
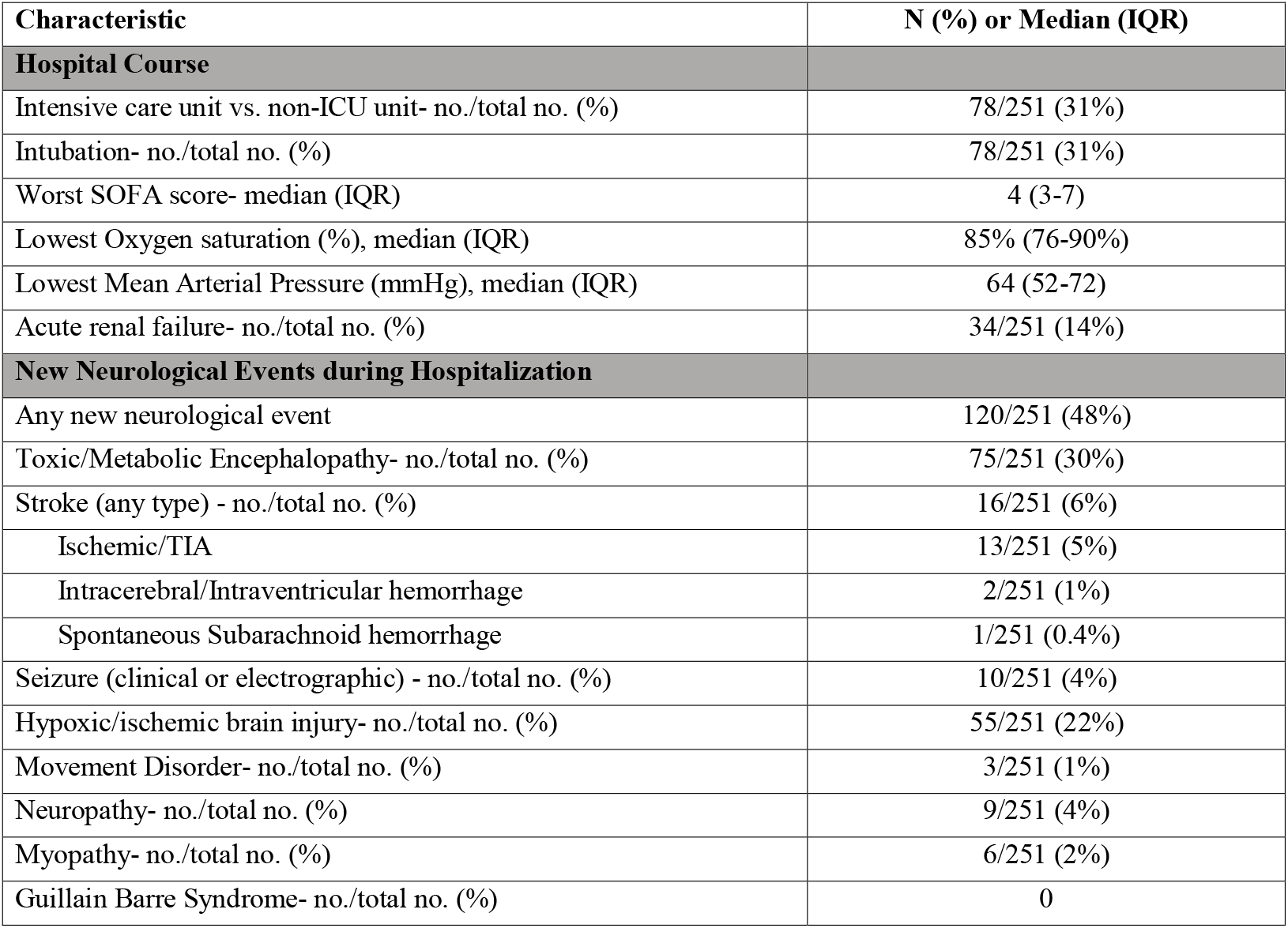
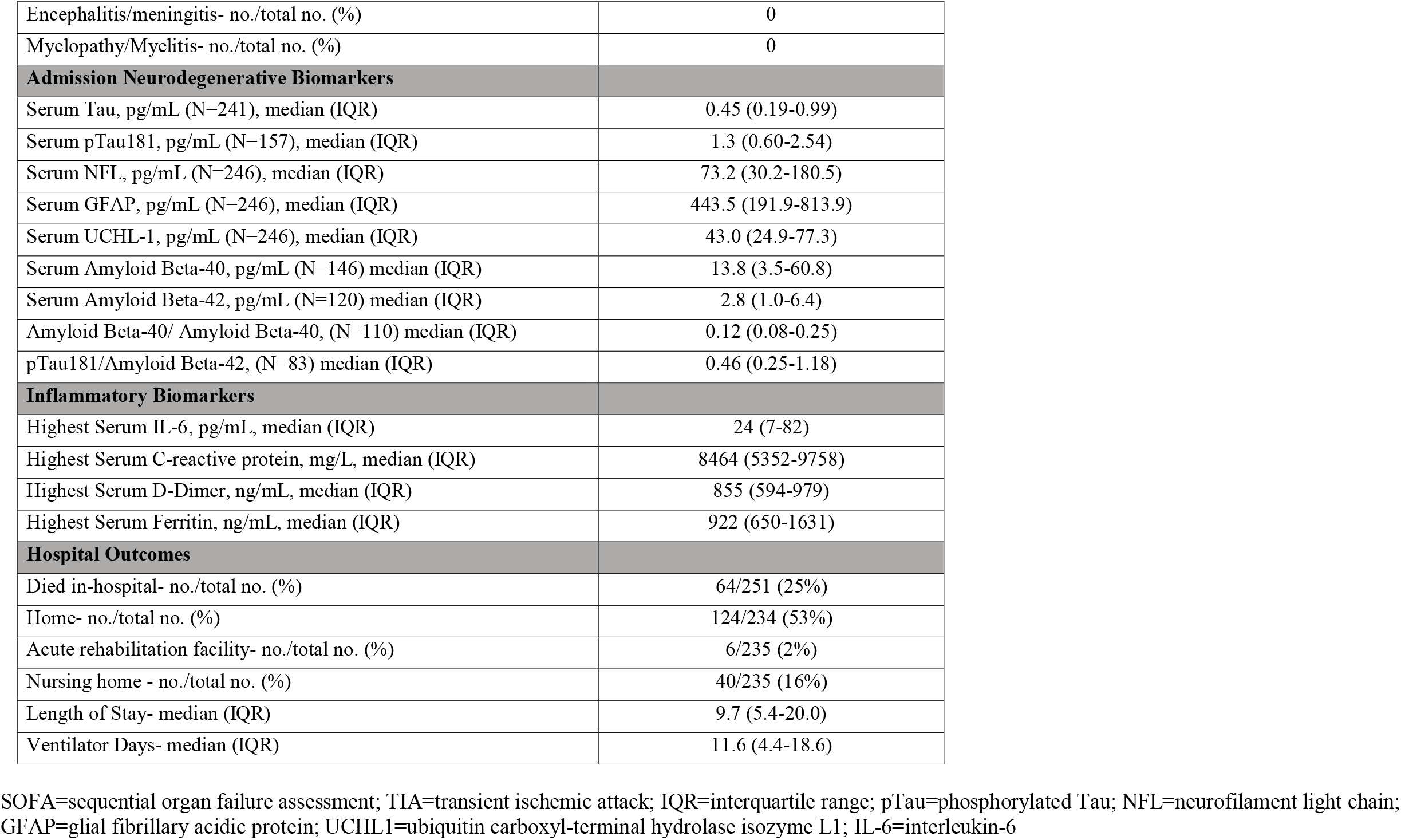
Characteristics of Hospitalized COVID-19 patients (N=251)

Elevations in neurodegenerative biomarkers in COVID-19 patients were significantly correlated with older age, and increased severity of illness (requirement of mechanical ventilation, worse SOFA scores, lower O_2_ saturations, lower mean arterial blood pressures; Table 2). Levels of total tau, pTau181 and NFL correlated most strongly with severity of COVID illness (green color on heat map indicates increasing Spearman’s rho correlation coefficients). Significant correlations were also identified between ptau-181, NFL, GFAP and elevated admission D-Dimer levels, though correlations were not observed with other inflammatory markers sampled at admission (IL-6, CRP, or ferritin; Table 2).

**Table 2.** Heat map of Spearman’s correlation coefficients among neurodegenerative biomarkers and demographics, severity of illness and inflammatory markers among hospitalized COVID-19 patients. Green color signifies stronger correlation and red signifies weaker.

|  | **Tau**<br>**N=241** | **pTau181**<br>**N=157** | **NFL**<br>**N=246** | **GFAP**<br>**N=246** | **UCHL1**<br>**N=246** | **Amyloid**<br>**Beta-40**<br>**N=146** | **Amyloid**<br>**Beta-42**<br>**N=120** |
| --- | --- | --- | --- | --- | --- | --- | --- |
| **Demographics** |  |  |  |  |  |  |  |
| Age | **0.213** | **0.367** | **0.273** | **0.435** | 0.084 | **0.294** | 0.14 |
| Male gender | 0.024 | 0.068 | 0.031 | 0.058 | 0.124 | 0.044 | 0.079 |
| Race (white vs. other) | **0.149** | 0.147 | 0.058 | 0.129 | 0.061 | 0.096 | 0.104 |
| **Severity of COVID-19 illness** |  |  |  |  |  |  |  |
| Intubation | **0.232** | 0.054 | **0.276** | 0.108 | **0.186** | 0.005 | 0.021 |
| Worse SOFA score | **0.345** | **0.261** | **0.461** | **0.25** | **0.313** | 0.13 | 0.138 |
| Lowest O2 saturation | **0.138** | 0.039 | **0.176** | 0.117 | 0.126 | 0.079 | 0.033 |
| Lowest Mean Arterial Blood Pressure | **0.312** | **0.256** | **0.385** | **0.178** | **0.271** | 0.075 | 0.065 |
| Hypoxic ischemic brain injury | **0.177** | **0.264** | **0.206** | **0.133** | 0.125 | 0.044 | 0.034 |
| Ventilator days | 0.215 | 0.279 | 0.085 | 0.141 | 0.099 | **0.586** | 0.352 |
| LOS | **0.135** | 0.046 | **0.291** | 0.101 | **0.193** | 0.046 | 0.009 |
| **Inflammatory Markers** |  |  |  |  |  |  |  |
| Admission IL-6 | -0.031 | 0.026 | 0.069 | 0.003 | 0.038 | 0.144 | 0.022 |
| Admission CRP | 0.006 | -0.017 | 0.044 | -0.059 | 0.003 | -0.096 | 0.056 |
| Admission ferritin | -0.015 | 0.026 | 0.023 | -0.005 | 0.002 | 0.113 | 0.02 |
| Admission D-dimer | -0.022 | **0.188** | **0.167** | **0.139** | 0.035 | -0.026 | -0.074 |

Green=stronger correlation; Red=weaker correlation; **Bold** indicates $P < 0.05$ . SOFA=sequential organ failure assessment; O2=oxygen; LOS=length of stay; Max=maximum recorded during hospitalization; IL-6=interleukin-6; CRP=C-reactive protein; pTau=phosphorylated Tau; NFL=neurofilament light chain; GFAP=glial fibrillary acidic protein; UCHL1=ubiquitin carboxyl-terminal hydrolase isozyme L1.

Compared to COVID-19 patients without any new neurological events, those with new neurological events during hospitalization had significant elevations in total tau (0.56 versus 0.33 pg/mL, P=0.009), ptau181 (1.58 versus 1.11 pg/mL, P=0.042), NFL (100.7 versus 61.8 pg/mL, P=0.006) and UCHL1 (47.9 versus 36.8 pg/mL, P=0.002) levels. Neurodegenerative biomarker levels for tau, ptau181, GFAP, and NFL were even higher among COVID-19 patients with TME compared to those without TME (Figure 2). Similarly, patients who died in-hospital had significant elevations in these biomarkers compared to those who survived, and patients who were discharged home had significantly lower levels than patients with other discharge dispositions (Figure 2). Aβ-40 and Aβ-42 did not have clear associations with TME, in-hospital death or discharge home, though the ratio of pTau181/Aβ42 was significantly associated with TME and in-hospital death, and Aβ42/40 was associated with discharge home.

**Figure 2.**
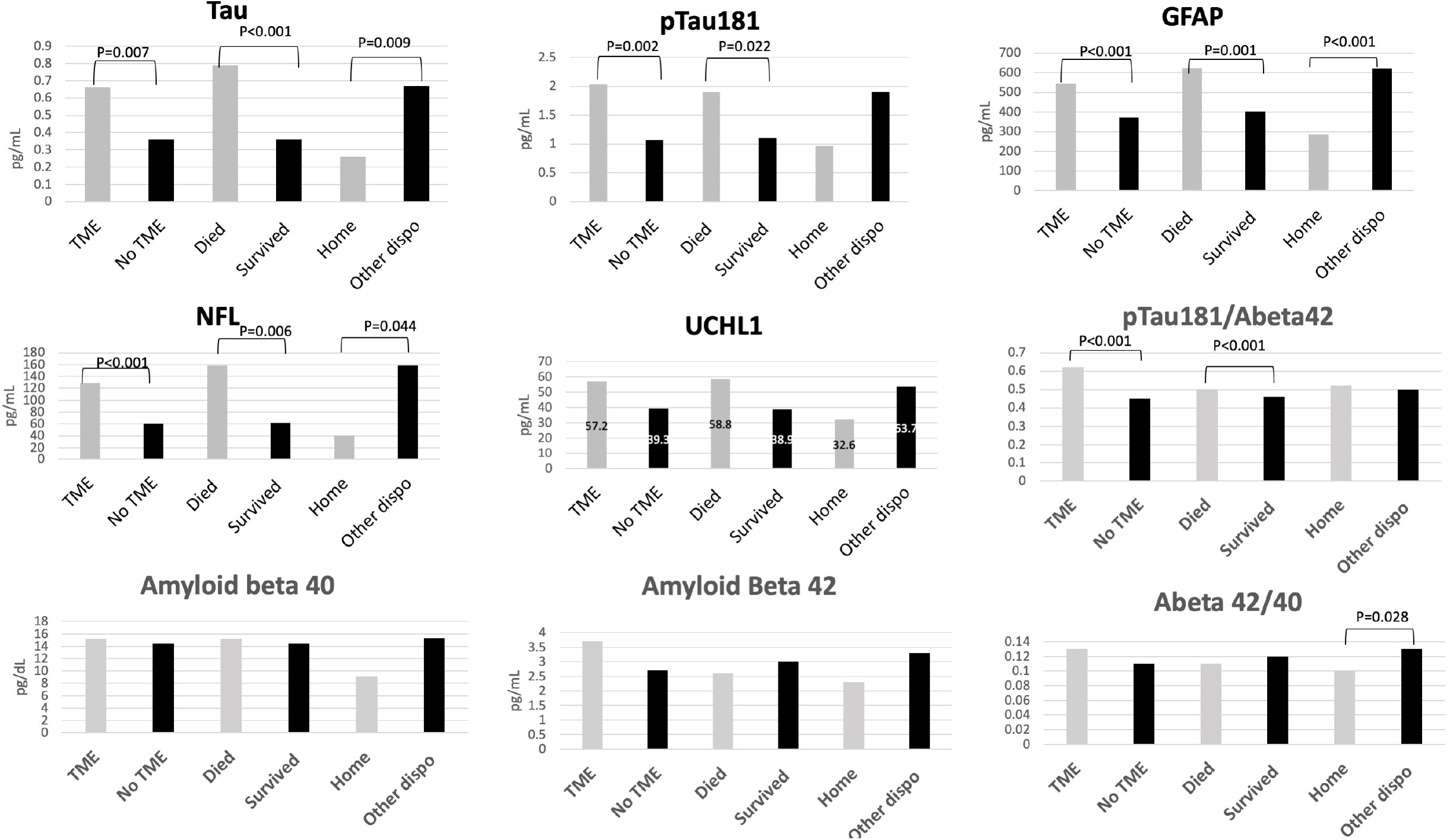
Serum neurodegenerative biomarkers in hospitalized COVID-19 patients (N=251) with and without Toxic-Metabolic Encephalopathy (TME), in-hospital death versus survival, and discharge home versus other discharge dispositions. pTau=phosphorylated Tau; NFL=neurofilament light chain; GFAP=glial fibrillary acidic protein; UCHL1=ubiquitin carboxyl-terminal hydrolase isozyme L1.

In multivariable Cox regression analysis adjusting for age, sex, race, history of neurological disease, admission sequential organ failure assessment (SOFA) score, and admission oxygen saturation, TME was significantly associated with increased admission pTau181 (Hazard ratio per 1 pg/ml increase [HR] 1.064, 95% CI 1.017-1.114, P=0.007) and UCHL1 (HR 1.001, 95% CI 1.000-1.002, P=0.037). In-hospital death was associated with elevated pTau181/Aβ-42 (HR 1.045, 95% CI 1.008-1.083, P=0.045). Higher levels of total tau (HR 0.686, 95% 0.519-0.906, P=0.008), NFL (HR 0.994, 95% CI 0.991-0.997, P<0.001), and GFAP (HR 0.999, 95% CI 0.999-1.000, P=0.012) were associated with lower rates of discharge home, while higher serum levels of Aβ-40 (HR 1.003, 95% CI 1.000-1.005, P=0.028) were associated with increased rates of discharge home (Table 3).

**Table 3.**
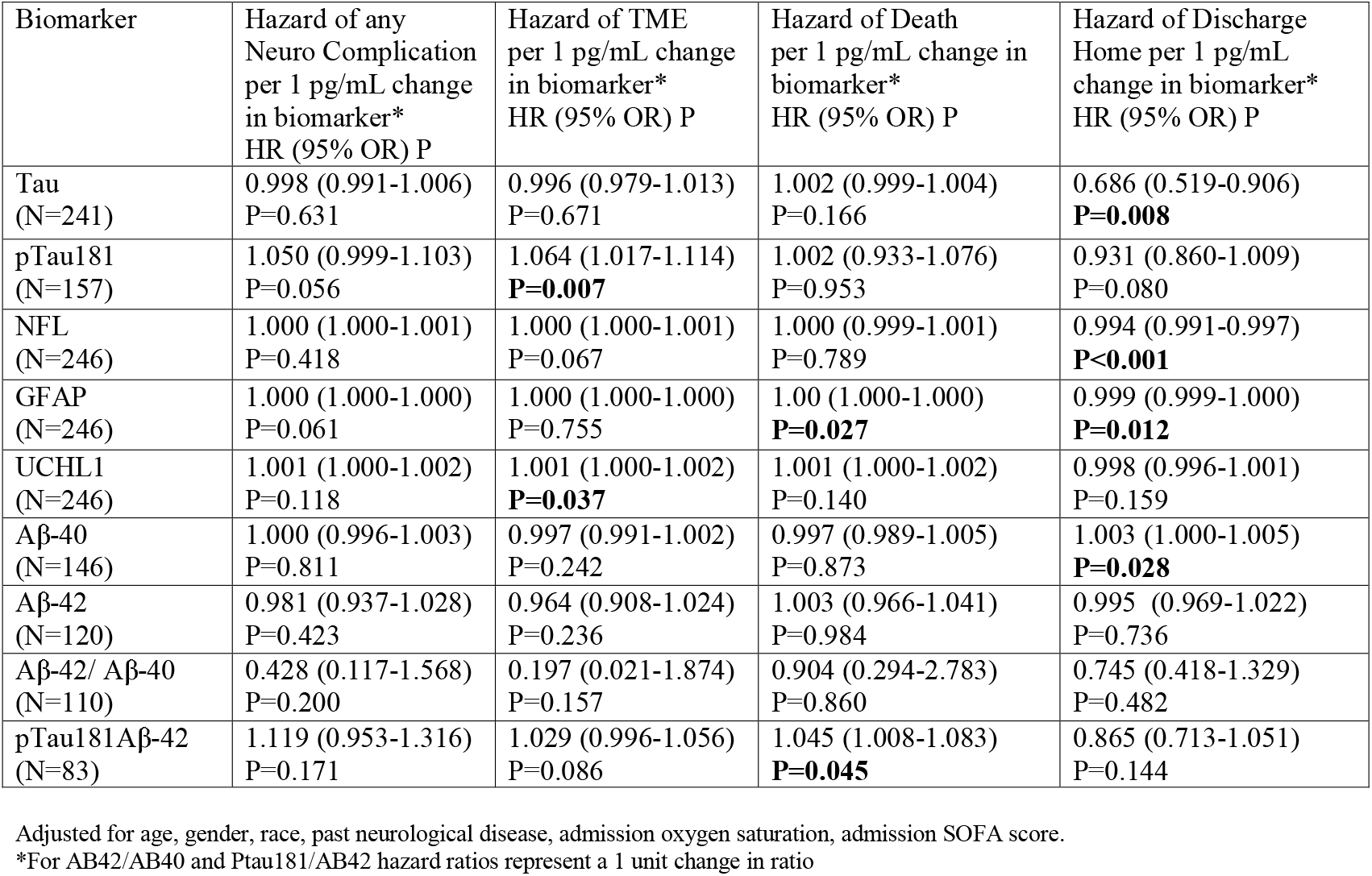
Multivariable Cox proportional hazard ratios among COVID-19 patients for each measured biomarker and the outcomes of: neurological events, toxic-metabolic encephalopathy (TME), in-hospital death and discharge home

| Biomarker | Hazard of any Neuro Complication per 1 pg/mL change in biomarker*<br>HR (95% OR) P | Hazard of TME per 1 pg/mL change in biomarker*<br>HR (95% OR) P | Hazard of Death per 1 pg/mL change in biomarker*<br>HR (95% OR) P | Hazard of Discharge Home per 1 pg/mL change in biomarker*<br>HR (95% OR) P |
| --- | --- | --- | --- | --- |
| Tau (N=241) | 0.998 (0.991-1.006)<br>P=0.631 | 0.996 (0.979-1.013)<br>P=0.671 | 1.002 (0.999-1.004)<br>P=0.166 | 0.686 (0.519-0.906)<br>**P=0.008** |
| pTau181 (N=157) | 1.050 (0.999-1.103)<br>P=0.056 | 1.064 (1.017-1.114)<br>**P=0.007** | 1.002 (0.933-1.076)<br>P=0.953 | 0.931 (0.860-1.009)<br>P=0.080 |
| NFL (N=246) | 1.000 (1.000-1.001)<br>P=0.418 | 1.000 (1.000-1.001)<br>P=0.067 | 1.000 (0.999-1.001)<br>P=0.789 | 0.994 (0.991-0.997)<br>**P<0.001** |
| GFAP (N=246) | 1.000 (1.000-1.000)<br>P=0.061 | 1.000 (1.000-1.000)<br>P=0.755 | 1.00 (1.000-1.000)<br>**P=0.027** | 0.999 (0.999-1.000)<br>**P=0.012** |
| UCHL1 (N=246) | 1.001 (1.000-1.002)<br>P=0.118 | 1.001 (1.000-1.002)<br>**P=0.037** | 1.001 (1.000-1.002)<br>P=0.140 | 0.998 (0.996-1.001)<br>P=0.159 |
| Aβ-40 (N=146) | 1.000 (0.996-1.003)<br>P=0.811 | 0.997 (0.991-1.002)<br>P=0.242 | 0.997 (0.989-1.005)<br>P=0.873 | 1.003 (1.000-1.005)<br>**P=0.028** |
| Aβ-42 (N=120) | 0.981 (0.937-1.028)<br>P=0.423 | 0.964 (0.908-1.024)<br>P=0.236 | 1.003 (0.966-1.041)<br>P=0.984 | 0.995 (0.969-1.022)<br>P=0.736 |
| Aβ-42/ Aβ-40 (N=110) | 0.428 (0.117-1.568)<br>P=0.200 | 0.197 (0.021-1.874)<br>P=0.157 | 0.904 (0.294-2.783)<br>P=0.860 | 0.745 (0.418-1.329)<br>P=0.482 |
| pTau181Aβ-42 (N=83) | 1.119 (0.953-1.316)<br>P=0.171 | 1.029 (0.996-1.056)<br>P=0.086 | 1.045 (1.008-1.083)<br>**P=0.045** | 0.865 (0.713-1.051)<br>P=0.144 |

Adjusted for age, gender, race, past neurological disease, admission oxygen saturation, admission SOFA score.
\*For AB42/AB40 and Ptau181/AB42 hazard ratios represent a 1 unit change in ratio

Compared to non-COVID controls, COVID-19 patients had significantly higher NFL and GFAP levels than non-COVID AD, MCI and normal controls, while UCHL1 levels were significantly higher in COVID patients compared to MCI and normal subjects, and were similar to levels observed in AD subjects (Figure 3).

**Figure 3.**
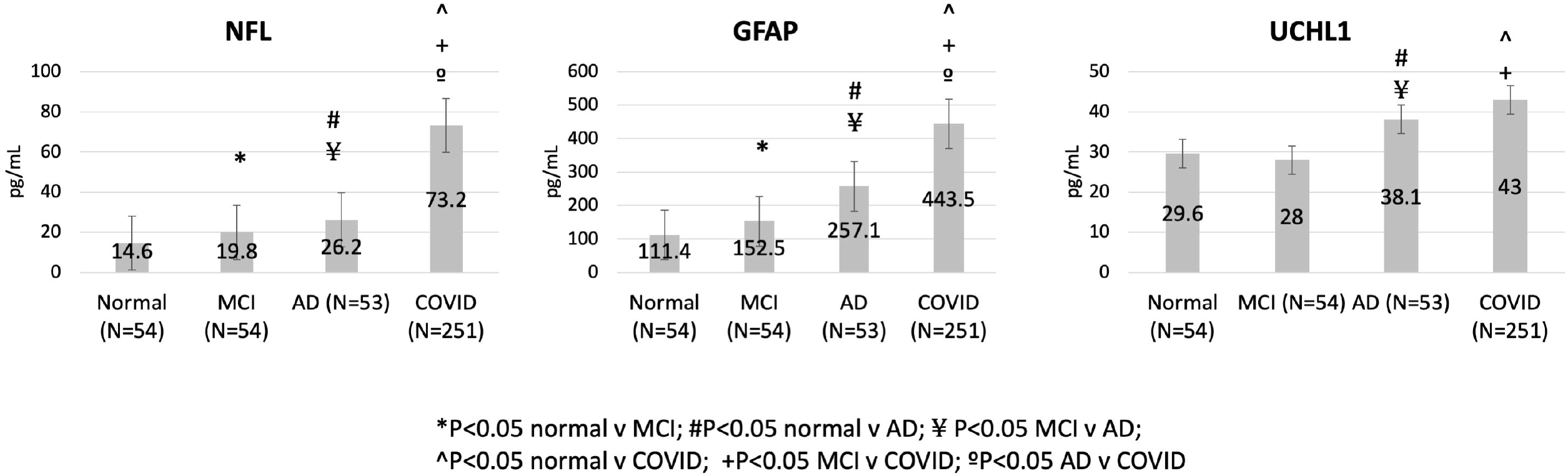
Plasma neurodegenerative biomarkers in controls (N=161 cognitively normal, mild cognitive impairment, and Alzheimer’s dementia patients) and serum biomarker levels in hospitalized COVID patients (N=251). MCI=mild cognitive impairment; AD=Alzheimer’s dementia; pTau=phosphorylated Tau; NFL=neurofilament light chain; GFAP=glial fibrillary acidic protein; UCHL1=ubiquitin carboxyl-terminal hydrolase isozyme L1; Abeta=Amyloid beta

## DISCUSSION

In this study we identified significant elevations in blood biomarkers of neuronal and glial degeneration among hospitalized COVID-19 patients with clinical signs of neurological injury, specifically TME. Indeed, higher admission levels of neuronal degeneration markers ptau181 and UCHL1 were significantly associated with TME, even after adjusting for age, sex, race, prior neurological disease and severity of COVID-19 illness. Similarly, in multivariable analyses, elevations in total tau, NFL, and GFAP, in particular, were associated with reduced likelihood of discharge home. Furthermore, we found that levels of NFL, GFAP and UCHL-1 were as high as, or significantly higher than those observed in non-COVID patients with AD, indicating a profound neurological insult in these patients. Strengths of this study include the prospective ascertainment of new acute neurological disorders among COVID-19 patients, inclusion of a variety of both neural and glial degenerative markers and AD-specific pTau181, use of blood rather than cerebrospinal fluid biomarkers (which makes this study feasible across a larger number of patients), and comparison to well characterized COVID-negative control groups with discrete levels of cognitive impairment.

We found significant correlations between neurodegenerative biomarkers and the inflammatory marker D-Dimer, which may provide some insight into mechanisms of acute brain injury following SARS-CoV-2 infection. Hypoxia and hyperinflammation, both hallmarks of acute COVID-19, have been linked to the development of AD-type pathology in non-COVID populations via upregulation of enzymes in the amyloidogenic pathway and downregulation of proteins that break down Amyloid β (28). Hypoxia-induced tau phosphorylation with corresponding memory deficits has been documented in animal models exposed to hypoxia for 6 hours/day for one to 8 weeks(29) and elevations of inflammatory cytokines such as IL-6 and IL-1 correlate with cognitive dysfunction and the promotion of amyloid plaque and neurofibrillary tangle pathology in animal models(30). We have previously found that COVID-19 related TME is significantly associated with elevations in inflammatory markers including IL-6, D-dimer, CRP, ferritin (22). IL-6, in particular, is known to promote endothelial dysfunction and vascular permeability and may play a role in blood brain barrier (BBB) dysfunction following SARS-CoV-2 infection (31). Neuropathological data among COVID-19 decedents have revealed evidence of hypoxic injury as well as endothelial inflammation, and BBB disruption (21, 32-35) that may be mediated, in part, by a COVID-related hyperinflammatory state. BBB disruption has also been implicated in AD-type pathology among non-COVID patients(36). Finally, another important association between COVID-19 and AD is the linkage to the apoE4 genotype, which is both a marker for increased COVID-19 severity(37, 38), and the most impactful genetic risk factor for late onset AD (39). We did not, however, identify correlates with blood Aβ-40 or Aβ-42 levels, which may indicate limitations in this assay or lack of a relationship in our cohort.

Elevations in blood and CSF NFL, GFAP and total tau among COVID-19 patients have been described by others, and in some cases, compared to normal control groups (4-6, 8, 9, 40-44). However, none of these studies explicitly excluded COVID-19 patients with a baseline history of dementia or cognitive decline, which likely would confound results. Additionally, NFL is not specific to the CNS and can be elevated in the context of peripheral neuropathy (45-47), including COVID-related critical illness neuropathy/myopathy (7) and Guillain-Barre Syndrome(47). Similarly, elevated GFAP has been reported in COVID-19 patients with critical illness neuropathy/myopathy and levels correlate with nerve amplitudes (7). More AD-specific biomarkers, such as pTau181 have not previously been explored in COVID-19 patients. In neuropathological studies of plasma pTau181 and NFL, both biomarkers accurately distinguish pathology confirmed AD from healthy controls, but only pTau181 distinguished AD from non-AD dementia cases and showed specificity for neuritic plaque pathology and Braak stage (14, 48). Similarly, pTau181 levels escalate progressively with worsening clinical dementia rating (CDR) scores and correlate with multiple cognitive domains, while NFL, total tau and amyloid ß levels do not perform as well(15, 49). We identified elevations across a spectrum of CNS specific markers, including neuronal (total tau, UCHL1), and astrocytic/glial markers (GFAP), as well as AD-related markers (pTau181).

There are limitations to this study. First, though we excluded patients with a history of dementia or cognitive decline, it is possible that some COVID-19 patients may have had preclinical or undiagnosed cognitive impairment. Second, biomarkers were only measured at one time point and we do not have data on trajectories of these markers. Some studies have identified persistent elevations in blood NFL levels for 30-50 days in small COVID-19 cohorts (8, 41), while GFAP levels may initially spike and then decline after the acute phase of infection(41). The association of these biomarkers with formal cognitive testing following the acute phase of COVID-19 has not been demonstrated and is an active area of research needed to unravel the long-term cognitive implications of elevated neurodegenerative biomarkers. Last, we compared serum biomarkers in COVID-19 patients to plasma levels in non-COVID controls, because pre-COVID banked serum specimens were not available in controls. Though NFL, GFAP and UCHL-1 levels are equivalent in serum and plasma(25), we were unable to compare total tau, ptau181 or Aβ levels to controls due to differences in specimen type(25, 26).

## CONCLUSIONS

Serum neuronal, glial and axonal neurodegenerative biomarkers, including total tau, pTau181, UCHL1, GFAP and NFL were significantly elevated in patients with encephalopathy and worse discharge disposition following hospitalization for COVID-19. These markers correlated with the severity of COVID illness. Furthermore, levels of NFL, GFAP and UCHL1 in hospitalized COVID patients were similar to, or higher than levels observed in non-COVID Alzheimer’s disease patients. Additional studies tracking trajectories of these biomarkers over time and their association with long-term cognitive outcomes among COVID-19 survivors are warranted.

## Data Availability

Data will be made available upon reasonable request

## ACKNOWLEDGEMENTS/CONFLICTS/FUNDING

We would like to thank the patients and families who participated in this study.

This study was funded by a grant from the NIH/NIA (PI: Wisniewski) COVID-19 administrative supplement 3P30AG066512-01

The authors report no relevant conflicts of interest.

